# Is execute attention affected by environmental lighting conditions?

**DOI:** 10.1101/2024.06.11.24308756

**Authors:** Nino Wessolowski, Raschida Jasmin Rahim

## Abstract

Bright daylight has various positive influences, such as the long-term synchronization of circadian rhythms and an associated alertness that creates optimal conditions for attentional performance. However, the short-term effects of light on attention have not yet been sufficiently researched. Studies of these short-term effects on selective attention also showed partly contradictory effects. To investigate these short-term effects, 95 participants completed the Attention Network Test, under conditions of bright daylight or warm-white light. The focus of the present study was on executive attention, because this construct comes closest to the ANT short form of selective attention used. A significant enhancement of executive attention was observed under bright daylight conditions. This specifically means a short term effect of bright daylight on executive attention and thus selective attention in young adults.

## Introduction

Natural sunlight affects humans, both physiologically and psychologically. Sunlight synchronizes the “internal clock,” influencing, among others, our sleep-wake cycle, mental energy, pleasure, motivation, and attention [1]. Therefore, artificial daylight is used indoors, to optimize room lighting or as a specific intervention in light therapy. Although field and laboratory studies have shown positive effects of intense artificial daylight on attention in different populations and under different conditions compared to neutral and warm white light and dimmed illuminance levels [2 - 9], not all investigations have yielded significant effects [9]

However, these empirical findings can only be explained theoretically to a limited extent, because the mechanism of light interventions during the day are still the subject of controversy [2,8]. Even those studies that point beneficial artificial light effects [3,7] have yielded heterogeneous results on individual subscales of the frequently administered d2 test of attention [10]: Increase in the speed-based total value (KL) vs a reduction in the error rate (F). Among the various possible causes underlying the diversity of extant findings, methodological differences in rather global forms of assessment of the construct of attention, particularly deserve closer consideration. The present study, therefore, investigates the effects of daylight on executive attention by employing an established and highly robust method resistant to confounders.

### 1.1 Pathways of the effect of light on human behavior

Regarding the general effects of light on human behavior, three different explanations are currently discussed [3,8].

(1. Visual pathway) In the retina light hits the retinal cones responsible for the perception of color and the retinal rods responsible for the perception of brightness; these impressions are then transmitted through the ganglion cells and through the optical nerve to reach the visual center in the occipital lobe, where a visual impression emerges [11]. An increase in illuminance results in an improvement in visual perception by means of increased brightness and color contrasts [12], which can in turn counteract visual impairments to some degree [13]. In addition, increased illuminance can also alleviate signs of fatigue as well as eye pain and headaches [12,13]. In the present study, this explanation is the visual pathway less possible. This is due to the absence of any change in the illuminance of the monitor, only that of the environmental condition. However, the improvement in vision by increasing the illuminance could be partly responsible for the effects of the reference studies that conducted paper-based attention tests.

(2. Neurobiological non-visual pathway) In addition to the retinal rods and cones, a third type of receptor has been identified at the retina, the intrinsically photosensitive retinal ganglion cells [14–16]. These receptors cover the retina like a spider web and are directly linked to the suprachiasmatic nucleus via the retinohypothalamic tract. This nucleus is the primary clock for all circadian body functions and controls these functions through the hormones melatonin and cortisol as well as the cryptchrome proteins CRY and PER [11]. Melatonin is produced in the pineal gland from the precursor serotonin and is suppressed by light. Cortisol is controlled indirectly through the release of adrenocorticotropic hormone, and its rhythm is roughly opposite that of melatonin release. Furthermore, the exact temporal course of light incidence on the retina, from changes in neuronal and hormonal activity to manifest changes in perception and behavior, is not yet fully clarified. In addition to the duration and intensity of exposure, individual and situational factors play a major factor [17,18].

(3. Psychological Pathway) Light may yield psychological effects by mechanisms of classical conditioning [19] and mood [17,18]. For example, dark-warm light like an open fire has been associated with cozy memories, yielding states of relaxation, etc. Dark and cold light, like in the cellar, is paired with unpleasant experiences and reminds one of fear or depression [19]. Bright daylight is linked to activity and creates a mood that increases concentration [17,18]. Knez [17], for example, showed a connection between light, mood and attention. The results of this study showed that a colour temperature which induced the least negative mood enhanced the performance in the long-term memory and problem-solving tasks, that require selective attention.

The mechanisms of the effect of light on immediate experiential and behavioral changes during the day are finally not clear. In addition to the duration and intensity of light exposure, individual and situational factors also interact in light application in everyday life: for example, age and gender [17,18] as well as anatomical characteristics [1], have an influence on the effect of light as individual factors. Situational factors of light applications include the location, occasion, and setting, as well as the form of the relationship with the people involved [20]. Output arousal may also play an important role in the effect of light on humans, especially in the stress response. A connection between light, attention and arousal was found [21,22].

Also the exact connection between light as a zeitgeber for circadian functions and immediate changes in short-term behavior during the day has not yet been completely clarified. However, the fact that short-term light interventions during the day also activate brain areas belonging to the circadian system [23] as well as studies have shown that light interventions during the day can influence alpha waves in the brain [24,25]. The research goal is to find further evidence at the behavioral level for the effect of light during the day.

### 1.2 Previous studies on the attentional effects of light

While there are fewer studies of high quality in adults, initial controlled studies on the use of artificial daylight in school settings showed that light supports attention in the age group of children and adolescents. For instance, Barkmann, Wessolowski, and Schulte-Markwort [3] examined the effect of artificial daylight on attention-related task performance in the setting of a field study in primary and secondary schools. Among other lighting conditions in variable light, a daylight condition (1060 lx and 6500 K) was compared with a control condition (300lx and 4000K). In a sample of 110 pupils, the concentration performance (CP; number of detected target items minus omission errors searching through lines of characters for prespecified target items) was measured using the d2 test of attention [10] and the ELFE 1-6 [21] as well as the LGVT 6-12 [22] reading tests. Under artificial daylight, there were significantly fewer errors (missed target items and false marked characters) in the d2 test of attention and significantly more words read in the reading tests. No effects were found in the lines processed per time unit (edited target items) and CP of the d2 test of attention.

A supplementary laboratory study by Wessolowski [26] showed comparable results in 95 adult students with significant effects of small size in overall error, omission errors and, contrary to the analogous field study, in working speed.

The first of three experiments of Sleegers, Moolenaar, Galetzka, Pruyn, Sarroukh and van der Zande [7] was a quasi-experimental field study with n = 98 students in grades 4 and 6 (M=10.0 years) from one elementary school. In total, the d2 test of attention was administered at three different time points (pre-measurement, first post-measurement, second post-measurement) under different light conditions in an intervention and a control group. The interval between pre- and post-measurements was one month, the interval between the two post-measurements one week. In the pre-measurement, the “standard” light program (300lx and 3000-4000K) was used for both groups. In the post-measurements, a daylight program (1000lx and 6500K) was used in the intervention group, while the “standard” light program was used in the control group. The intervention group scored significant better (large effect sizes) on d2 overall error and CP compared to the control group. In the second experiment of Sleegers et al., n = 37 students at the age of 10 years from two schools were examined with otherwise the same procedure, light conditions and measuring instruments as in the first experiment. The results of this second experiment largely replicated those of the first experiment. The third experiment realized a randomized design in a darkened lecture room with a total of n = 55 students from six schools. No significant effects were found in terms of overall error, speed or CP.

The aim of the study by Auras, Barkmann, Niemeyer, Schulte-Markwort and Wessolowski [2] was to test specific light interventions on attention in the Child and Adolescent Psychiatry. Using a quasi-experimental A-B-A-B study design, a daylight programme (A) with high illuminance (5312 K, 793 lx) was tested against a comparative light programme (B; 4081 K, 378 lx). CP, overall errors and working speed of 30 patients (age M = 11.5 years) was assessed using the d2 test of attention of [10] and subjective self-assessment for attention and stress level was recorded using questionnaires. In the d2 test of attention there were significant improvements of medium size under the daylight programme condition in CP and working speed but there were no effects in overall errors. The subjective self-assessments, on the other hand, showed a decrease in concentration and an increase in stress and fatigue under daylight conditions.

Weitbrecht, Bärwolff, Lischke and Jünger [9] investigated the influence of the correlated color temperature of light on concentration and creativity in n = 50 students and employees. The three light scenarios (3000 K, 4500 K, 6000 K) were given with a constant illuminance of 1000 lx in a darkened room by an LED surface area light. Among others, the d2-bq test (short form of d2 test of attention), a creativity test [28], a word test (in which words of a certain category must be recognized in a box with letters) [29] and a logic test [28] were used. The order of the three light scenarios was randomized. In the d2-bq test, no significant differences were found between the light conditions. Under the warm light condition (3000 K), improvements in the creativity test were observed compared to the conditions of 4500 K and 6000 K. Significant advantages in favor of the daylight condition (6000 K) were found in the word test, which also requires attention. In the logic test (also attributed to creativity), the highest performance was achieved at 3000 K compared to 4500 K and 6000 K. Accordingly, the authors conclude that performance improvements were significant in terms of creativity in warm light (3000 K) and in terms of attention in light with higher blue content (4500 K, 6000 K), even though their results failed to reach the level of significance.

In summary, five of six studies using the d2 test of attention found significant effects when comparing performance under conditions of bright daylight. But these findings are appear somehow inconsistencies. While Sleegers et al. [7] found effects both in CP and in the decrease of overall errors, Barkmann et al. [3] only found effects in the decrease of overall errors, Wessolowski [26] found effects in overall errors and working speed and Auras et al. [2] found an increase in CP and no decrease of overall errors.

It should be noted, however, that the performance measures provided by the d2 test are unspecific with regard to the underlying attentional processes and were observed in children and adolescents.

### 1.4 Questions and hypotheses of the present study

The results of controlled light studies in children and adolescents speak for an effect of daylight on attention. However, participants subjectively report more stress [2] and the objective effects are found on different scales of the d2 attention test. Perhaps because the increase in performance due to daylight is at the expense of the perception of stress? Furthermore, controlled light studies in the age group of adults are lacking. Therefore, the present study is intended to clarify more precisely the impact of light on young adults. In addition, this study aims to understand the effect of light on the process of executive attention. Therefore, we analyzed executive attention with the Attention Network Test (ANT) [30] in short form more elaborately than was previously the case with the d2 test of attention. The d2 test measures rather broadly and less process-specific. This could also explain the heterogeneous d2 findings of the previous studies. We assume that congruency (executive Attention) in the ANT comes closest to the construct of selective attention in the d2 test of attention. The ANT combines two of the classical paradigms of visual attention (i.e., the Posner and Eriksen paradigms). In addition to Executive Function, the ANT incorporates the Orienting and Alerting networks. In principle, light effects can also be expected for these factors [31,32]; however, due to the short duration of the experiment [30], it is possible that these effects can only be detected minimally or not at all. Fan et al. [30] demonstrated that an enhancement in the performance of executive attention tasks correlates with the activation of the anterior cingulate cortex (ACC) in the human brain. If a discernible light effect is observed in the current experiment concerning executive function, this would provide additional evidence for the biologically non-visual function in short-term light effects and further support for the short-term activation of the circadian system, as both the SCN and ACC are constituents of the limbic system [11]. Based on observations during previous studies [2], we also explicitly surveyed subjective stress perception during light exposure using questionnaires. This is important because stress affect performance. The Yerkes and Dodson law shows an inverted u-shaped relationship. This implies that performance initially rises in proportion to stress, and above a medium level of stress, performance reaches its best value; if stress now continues to rise, this increasingly leads to losses [27].

In sum, the present study aimed to answer the following three questions: 1. Are higher performances in executive attention observed under bright daylight than under dimmed warm light conditions? 2. What effects can be observed in relation to orienting and alerting? 3. Are the two lighting conditions associated with different levels of subjective stress.

## Materials and Methods

### 2.1 Sample

The sample size was determined by an a-priori power analysis. Using G-Power [33], this showed an optimum sample size of n = 788 with expected small effects and p < .05. Since such a large sample has to be collected over several years in the complex laboratory experiment, the present study monitors the effects after data collection in the first semester.

A sample was recruited from Bachelor and Master students, their families and friends. All n = 91 participants were included in all analyses because there were no missing data. The large proportion of psychology students (75.8 %) and the high proportion of women (72.6 %) should be accentuated (see Table 1). None of the control variables showed any significant correlations with the ANT Outcome (target variables), so that these variables were not included as covariates in the analyses.

**Table 1.** Sample information.

|  | BL | WL | total | Significance between groups |
| --- | --- | --- | --- | --- |
| n (%) | 53<br>(58.2) | 38<br>(41.2) | 91<br>(100.0) | $X^2 = 2.473$, $df = 1$, $p = .116$ |
| age in years M<br>(SD) | 22.4<br>(1.77) | 23.3<br>(2.52) | 22.7<br>(2.12) | $T = 1.933$, $df = 1$, $p = .056$, $d = .411$, $CI: -0.011 - 0.831$ |
| sex f/m in % | 76.8/23.2 | 66.7/33.3 | 72.6/27.4 | $X^2 = 0.552$, $df = 1$, $p = .457$ |
| Students of<br>psychology in % | 69.2 | 80.4 | 75.8 | $X^2 = 4.526$, $df = 1$, $p = .033^*$ |

Notes. BL = bright daylight, WL = dim warm light, T = T-Test (two tailed), X2 = Chi-Square Distribution.

### 2.1 Design and Materials

The study took place in the laboratory of the Medical School Hamburg (MSH) in four identical sound insulated and ventilated test cabins (180 cm long and 220 cm high, light grey inner walls) (see Figure 1). A 72 x 35 cm cabin window was located behind the participants (left of the door). The cabin windows were facing away from the windows of the laboratory room (north-east), so that barely any natural daylight (see control variables) would enter the cabins (maximum value determined over all measurements: 30 lx). The cabins are each equipped with two chairs, one table, one computer, one screen, and two lamps.

**Figure 1:**
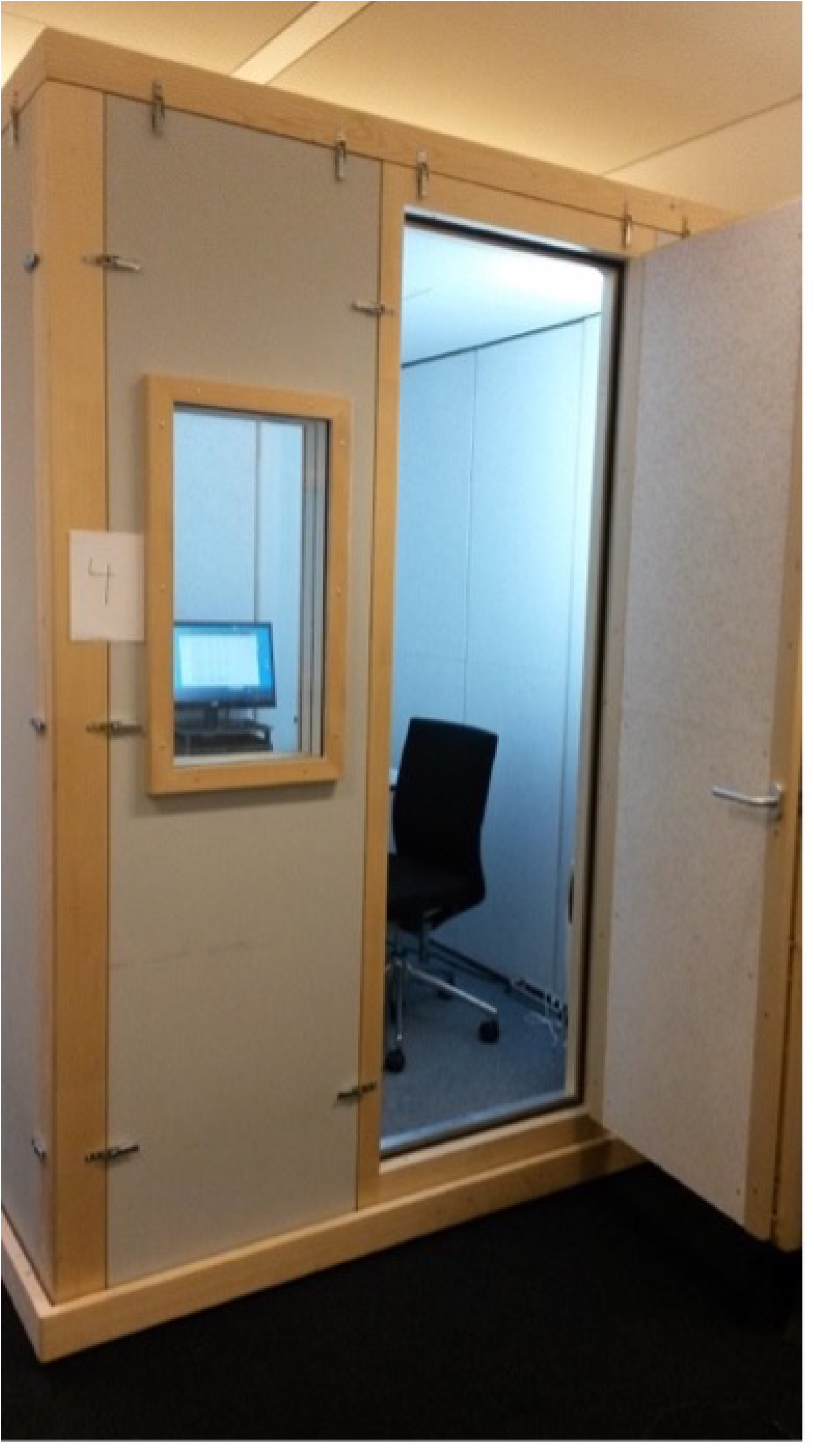
Cabin in the laboratory of the Medical School Hamburg (Picture by Johanna Hänsel).

The hardware was a Lenovo PC (Intel (R) Pentium (R) CPU G 3220 @ 3.00 Ghz, RAM: 4.0 GB) with Windows 7 Professional (64 Bit) and a computer monitor with a 22" LCD screen. The ANT (see below) was controlled and presented with the experimental software E-Prime 2.0 (Psychology Software Tools, Inc.).

As illustrated in Figures 1-3, the lighting for the experimental condition bright daylight was provided by two off-the shelf floor lamps (Ikea Not) which were equipped with two compact fluorescent lamps (one Philips T65 Softone Cool Daylight E27 20W 865 and one Philips Genie Cool Daylight E14 8W 865) and a surface mounted light (Luxero T5LL-21 W) with a fluorescent tube (FSL 21W 865/0). The light for the control condition dim warm-white was operationalized by the lefthand floor lamps (see Figure 3). This condition is equipped with another compact fluorescent lamp (Philips Tornado E14 12W 827). The movable arms of the floor lamps with the E14 sockets were fixed to the cabin interior in both conditions with the light cone aligned to the ceiling. The operationalization of the lighting conditions is discussed in 4.3 Limitations.

The cabins were equipped with a measuring device ALP-01 from Asensetek Lighting Passport. The illuminance and color temperature were measured horizontally on the work surface. Under the bright daylight (BL) experimental condition, an average of 517.0 lx / 5417 K was measured. This generated a condition with high illuminance and daylight color as well as a correspondingly high blue component (see Figure 2). An average of 67.5 lx / 2628 K was measured under the dim warm-white (WL), control condition resulting in low illuminance and warm-white color temperature with a correspondingly high red component (see Figure 3).

**Figure 2:**
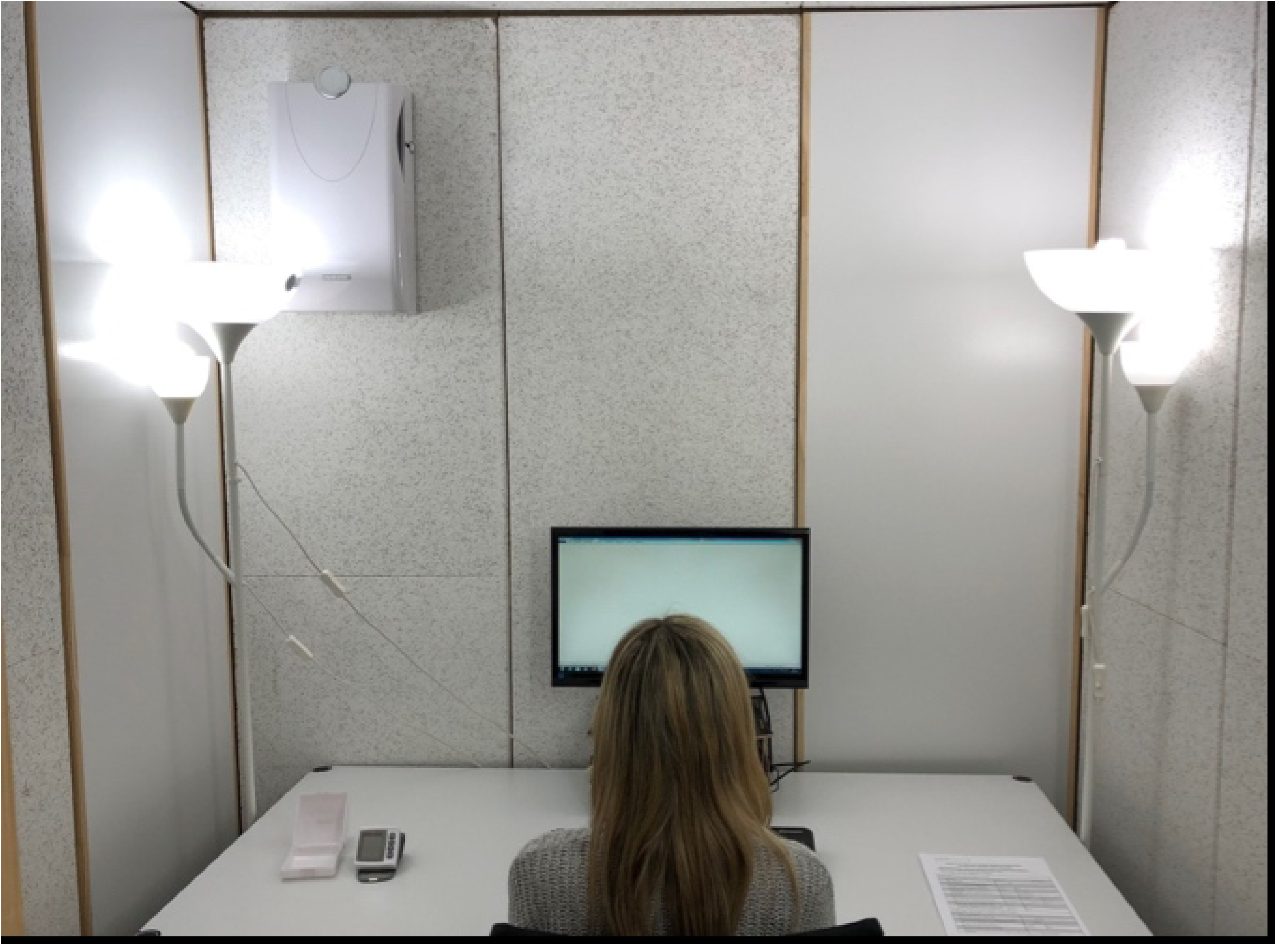
Cabin in the laboratory with experimental condition bright daylight (Picture by Nilay Dermidal and Ricardo Frink).

**Figure 3:**
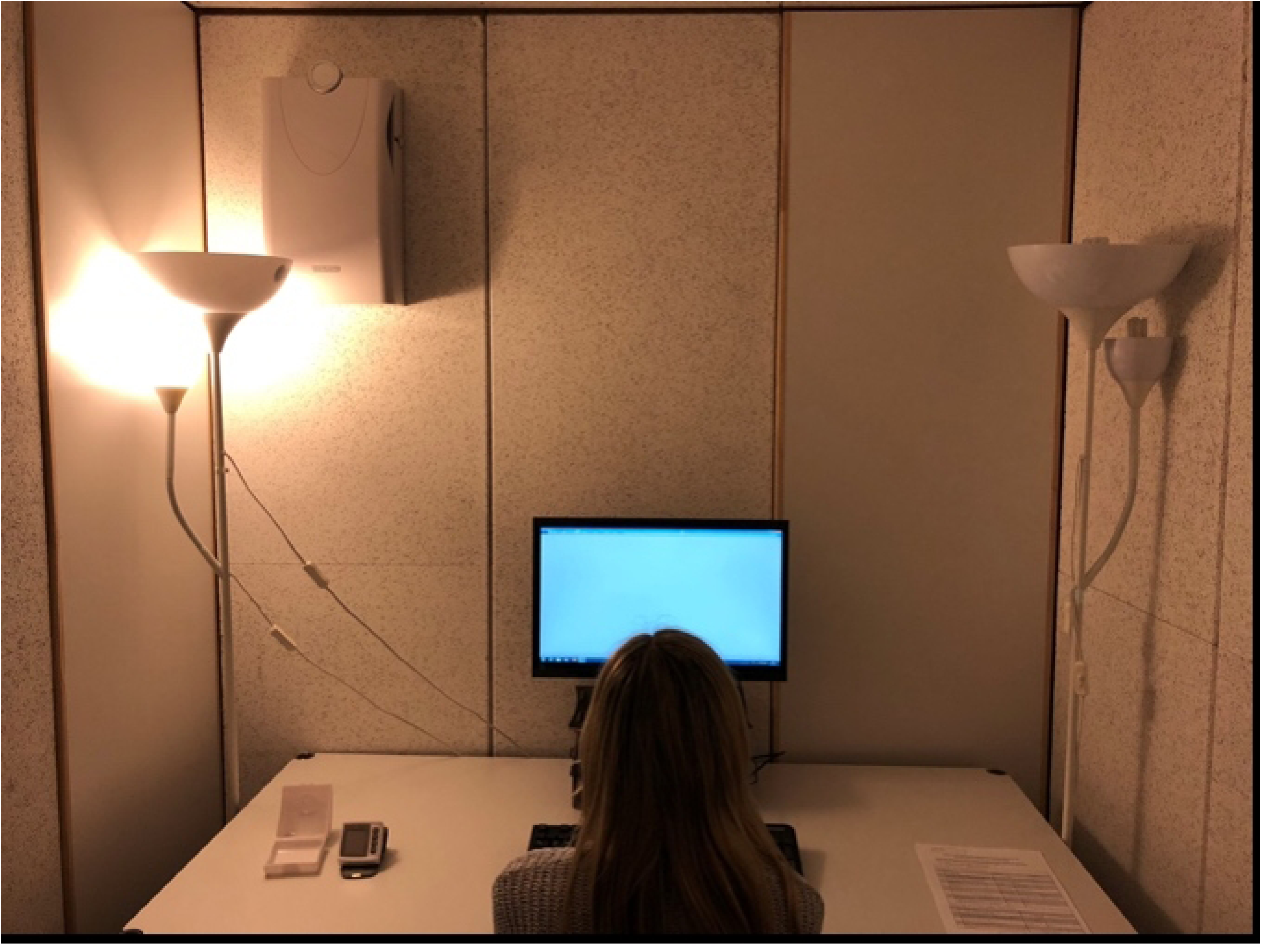
Cabin in the laboratory with control condition dim warm light (Picture by Nilay Dermidal and Ricardo Frink).

### 2.2 Dependent Variables

In this study the ANT-Short was used [30]. Participants were instructed to react as quickly as possible to the direction of a middle arrow on a computer screen and click with the left or right thumb on the left or right mouse button.

The Conflict effect (**Executive attention**) is calculated by the difference in reaction times between trials with congruent arrows and incongruent arrows [30]. The stimuli are illustrated in Figure 4.

**Figure 4.**
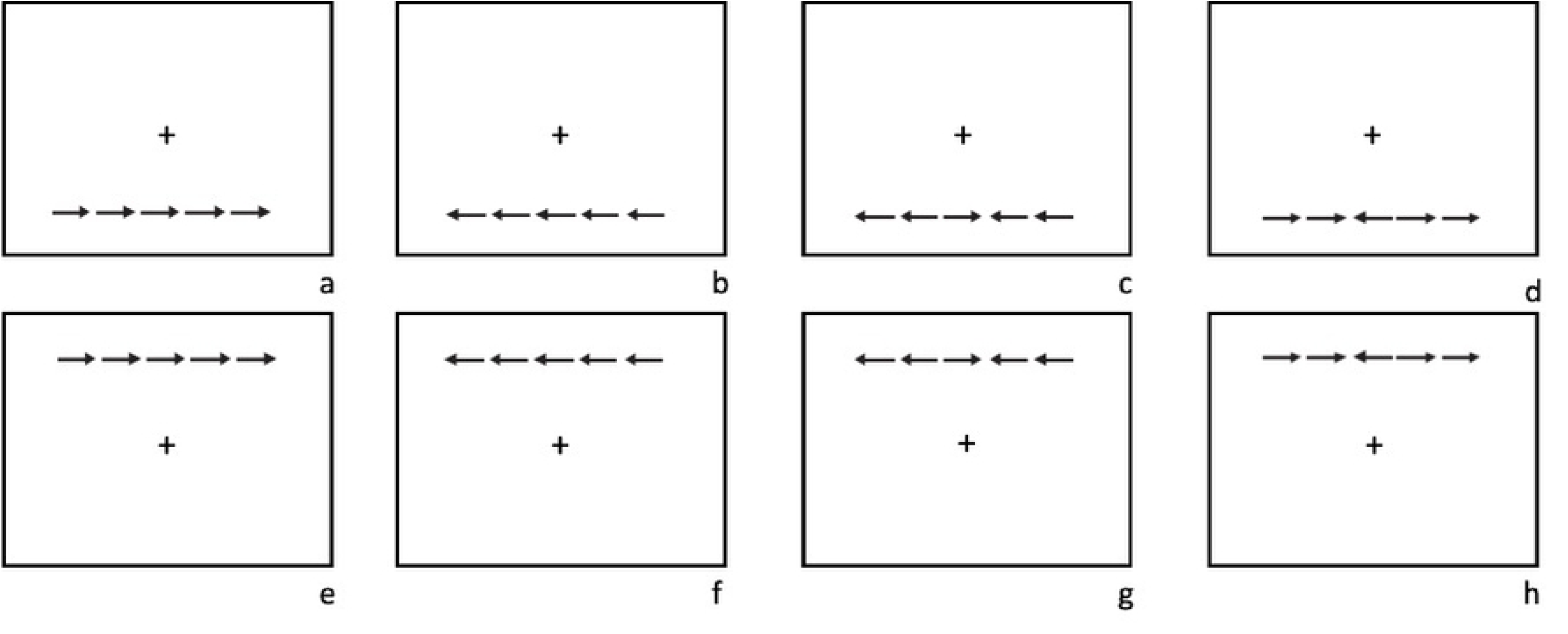
(a-h): congruent bottom (a-b), incongruent bottom (c-d), congruent top (e-f), incongruent top (g-h).

The **Orienting** effect is calculated by the difference in reaction times between trials with a center cue and spatial cue [30]. The stimuli are illustrated in Figure 5.

**Figure 5.**
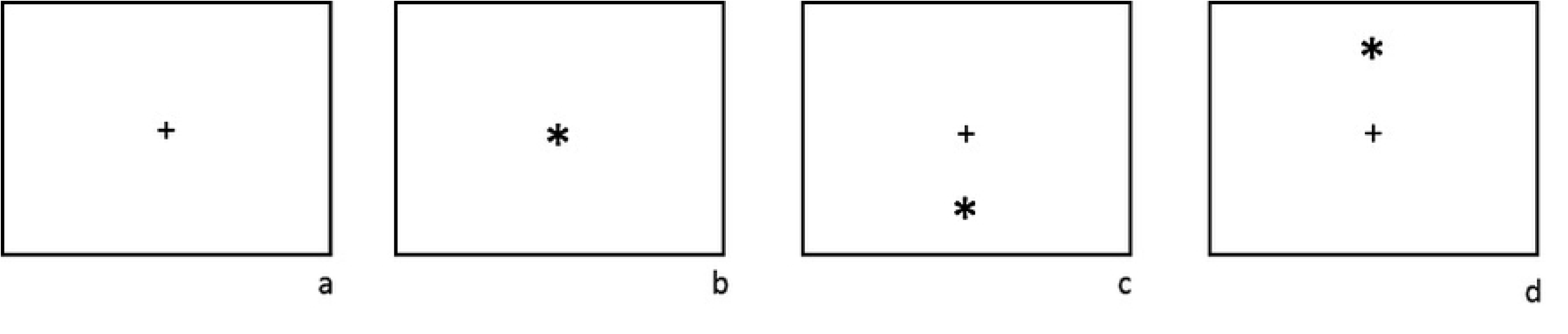
(a-d): no cue – only fixation cross (a), center cue (b), spatial cue bottom (c), spatial cue top (d).

The **Alerting** effect is calculated by the difference in reaction times between trials with no cue and a center cue [30]. The stimuli are also illustrated in Figure 5.

Based on the findings in the previous studies, ratings of **acute stress** were also obtained. The scale rest and restlessness of the multidimensional state questionnaire (MBDF) [34]was used. Self-formulated short scales were used to determine the following control variables: long-term stress, consumption of caffeine intake, chronotype, night sleep, nationality, mother tongue, glasses/contact lenses, left/right-handedness, and chronic diseases. In further details sunshine hours duration of the day [35], room temperature (temperature), date and time were recorded. Additionally, pulse and blood pressure were measured with blood pressure monitors (Sanitas SBC 15).

### 2.4 Procedure

The data collection took place from 20.04.2021 to 16.05.2021. The light condition was determined at random (coin toss). Accordingly, the luminaires were therefore fitted with the appropriate lamps for lighting condition, switched on and the room temperature measured. To ensure that the initial conditions were as comparable as possible and to allow the light exposure to take effect. The adult participants were informed in both written and verbal form about the study and gave their written consent. The participants were first asked to complete questionnaires. This should result in a total acclimatization and exposure of about 10 minutes before the actual experiment. The experimenter stayed in the cabin during the instruction of the ANT and the test trial. Afterwards pulse and blood pressure were measured. Then the experimenter left the cabin and the participants completed the experiment, consisting of three ANT blocks each containing 48 trials. Each of these blocks was followed by a response on the scale of calm / restlessness. Pulse and blood pressure values were measured for the second time and afterwards the participant was send-off.

### 2.5 Ethics

The experiment was conducted in accordance with the Declaration of Helsinki and was approved by the Ethics Committee of Medical School Hamburg (**MSH-2020105)**.

### 2.6 Statistics

An analysis of variance (ANOVA between subjects) was used. A Huynh-Feldt correction was used when the sphericity assumption was violated. The dependent variables were successfully tested for normal distribution by testing kurtosis and skewness.

## Results

### 3.1 Analysis of the ANT

Table 2 shows a significant effect of small size in the main variable of interest, executive function, in favor of the bright daylight group. This means that an average participant in the bright light group has a 11.3% shorter time difference between the congruence and no congruence conditions than an average participant in the control group with dimmed warm white light. The small effect for orienting (n.s.) and the absence of an effect for alerting are as expected.

**Table 2.** Analysis of the ANT (reaction time).

| | n | BL M (S) | WL M (S) | F | df | p | $\eta_p^2$ |
| --- | --- | --- | --- | --- | --- | --- | --- |
| Executive attention | 91 | 89.7 (0.11) | 101.2 (26.60) | 3.990 | 1 | .049* | 0.043 |
| Orienting | 91 | 56.6 (28.79) | 65.5 (27.60) | 2.108 | 1 | .150 | 0.023 |
| Alerting | 91 | 22.1 (21.04) | 19.6 (22.79) | 0.258 | 1 | .613 | 0.003 |

Notes. BL = bright daylight, WL = dim warm light, F = F-Value of ANOVA (two tailed), $\eta_p^2$ = Partital Eta Squared.

### 3.2 Stress

Table 3 shows that there was no significant light effect (time*group) on the perceived stress of the participants during the test. In contrast, a significant effect of small size was found for the time course.

**Table 3.**
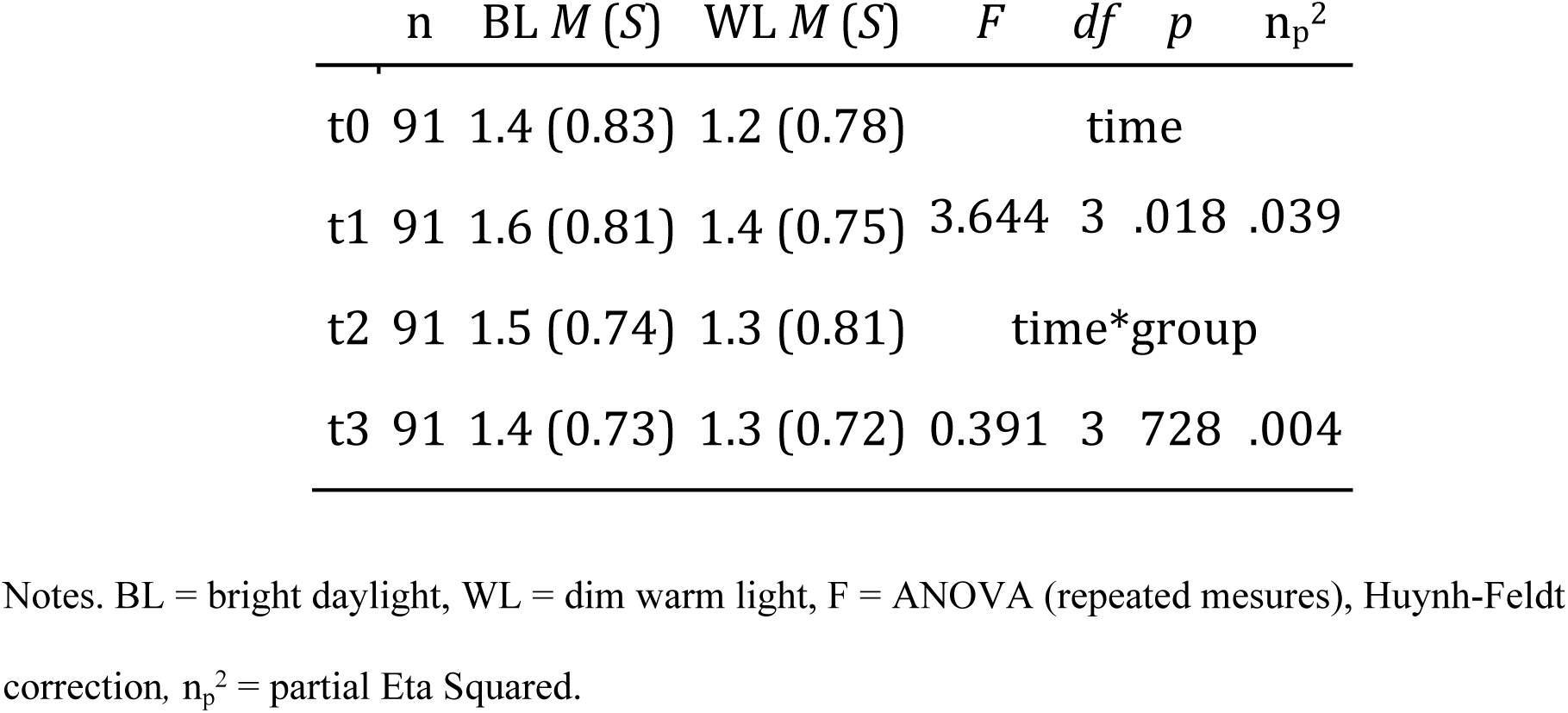
Analysis of perceived stress.

## 4. Discussion

### 4.1 Main Results

The present study investigates whether higher performances observed in attentional functions under bright daylight than under dim warm light and whether stress perception is different between the light conditions. The anticipated impact on executive function in favor of the bright daylight condition, compared to the dimmed warm white light condition, can be confirmed. As anticipated, the orienting effect is smaller in this experiment, and no alerting effect is observed. No light effect was observed in relation to stress.

### 4.2 The relationship of the present results with previous studies and the pathways of the effect of light on human behavior

The observations of Auras et al. [2] that daylight interventions subjectively lead to higher stress levels could not be confirmed in this study. No effect in the stress load was found. Presumably, the effect of light on the perception of stress depends on the state of arousal. Highly stressed people like the children in the psychiatric hospital in the study by Auras et al. [2] may first need a calming dimmed warm white light. However, the setting and the experimental population showed considerable differences between the present study and the reference study, so that further research on this question is necessary.

### 4.3 Limitations

In contrast to the preliminary studies, this study administered a computer-based test, whereas the reference studies are based on paper pencil tests [3,7,23]. In addition, the monitors emitted white light as screen background regardless of the lighting condition administered. So the contrast between the light conditions was not as pronounced as in the reference studies. In addition, the daylight condition in the reference studies by Barkmann et al. [3] and Slegers et al. [7] were approximately twice as intense with approximately 1000 Lux. In the study of Auras et al. [6], the daylight condition was also significantly brighter (793 lux) as was the case with Wessolowski [23], with almost 1300 lx. This could have resulted in possible effects of the light that could not be represented reliably. Another limitation is that the lighting conditions were not generated from only one luminaire as in Barkmann et al. [3] and Slegers et al. [7] but from different luminaires. Thus, other lighting parameters varied except for illuminance and the most similar color temperature. However, the present study was carried out entirely without funding from the lighting industry and was financed entirely from the authors’ private funds. This is certainly not ideal for an experiment, but the change of the lighting situation by different favorable light sources corresponds to frequent practice in real life.

The data collection took place in spring and early summer. Although the test cabins were largely isolated from natural daylight, the participants were certainly influenced by natural daylight experienced previously outside the lab. It is conceivable that natural daylight outside the experiment was much more effective than artificial light in the experiment and caused strong undesired stimulation, especially in the control condition. For example, Kent et al. demonstrated a significant correlation between natural daylight exposure of the participants, as measured by NASA satellites, and their performance on cognitive tests [36]. Although an influence of sunshine duration (local weather data) on the test results could not be determined (not shown here), it seems conceivable that our results are conflated by previous exposure to bright daylight.

### 4.4 Summary

In terms of executive attention in the ANT test utilized, the results favored the daylight condition. Since the test was computerized for both the daylight and warm white light groups with constant monitor settings, the effect cannot be solely attributed to the visual impact of light. Light seems to directly affect executive attention and presumably selective attention, with activation observed in the anterior cingulate cortex (ACC) [37], a component of the circadian system. Van der Valle also demonstrates that short-term light interventions result in the activation of brain regions associated with the circadian system [23]. This suggests that the short-term impact of light on attentional performance may share a similar biological, non-visual pathway with the circadian effect of light.

A non-significant effect of small magnitude was observed for orienting, and no effect was found for alerting, potentially attributable to the brief duration of the experiment. In principle, the experiment should be expanded on a larger scale and with more pronounced light contrasts. The lighting scenarios do not appear to influence subjects’ stress perception, at least not in this experiment; however, this could be attributed to sample size, experimental setting, and participants’ arousal levels.

## Data Availability

All relevant data are within the manuscript and its Supporting Information files.

